# Chronic post-COVID neuropsychiatric symptoms (PCNPS) persisting beyond one year from being infected during the ‘Omicron wave’

**DOI:** 10.1101/2024.09.01.24312691

**Authors:** Steven Wai Ho Chau, Timothy Mitchell Chue, Tsz Ching Lam, Yee Lok Lai, Rachel Ngan Yin Chan, Paul WC Wong, Shirley Xin Li, Yaping Liu, Joey Wing Yan Chan, Paul Kay-sheung Chan, Christopher Koon-Chi Lai, Thomas WH Leung, Yun Kwok Wing

**Affiliations:** Department of Psychiatry, Faculty of Medicine, The Chinese University of Hong Kong, Hong Kong SAR; Li Chiu Kong Family Sleep Assessment Unit, Department of Psychiatry, Faculty of Medicine, The Chinese University of Hong Kong, Hong Kong SAR; Department of Social Work and Social Administration, Faculty of Social Sciences, The University of Hong Kong, Hong Kong SAR; Department of Psychology, Faculty of Social Science, The University of Hong Kong, Hong Kong SAR; Center for Sleep and Circadian Medicine, The Affiliated Brain Hospital, Guangzhou Medical University, Guangzhou, Guangdong, China; Department of Microbiology, Faculty of Medicine, The Chinese University of Hong Kong, Hong Kong SAR; S.H. Ho Research Centre for Infectious Diseases, Faculty of Medicine, The Chinese University of Hong Kong; Division of Neurology, Department of Medicine and Therapeutics, Faculty of Medicine, The Chinese University of Hong Kong, Hong Kong SAR; Li Ka Shing Institute of Health Sciences, Faculty of Medicine, The Chinese University of Hong Kong, Shatin, Hong Kong SAR, China

## Abstract

The heterogeneity of chronic post-COVID neuropsychiatric symptoms (PCNPS), especially after infection by the Omicron strain, has not been adequately explored. Our pre-registered hypotheses are 1. chronic PCNPS in patients infected with SARS-CoV-2 over a year ago during the ‘Omicron wave’ showed a similar clustering pattern with symptoms in patients infected with pre-Omicron strain; 2. these chronic PCNPS are associated with a) clinical risk factors, such as, severity of the acute infection; b) socioeconomic status e.g., level of deprivation; and c) pre-infection vaccination status. We assessed 1205 subjects using app-based questionnaires and cognitive tasks. Partial network analysis on chronic PCNPS in this cohort produced two major symptom clusters (cognitive complaint-fatigue cluster and anxiety-depression symptoms cluster) and a minor headache-dizziness symptoms cluster, like our pre-Omicron cohort. Subjects with high number of symptoms (4 or more) can be further grouped into two distinct phenotypes: a cognitive complaint-fatigue predominant phenotype (CF) and another with symptoms across multiple clusters (AD-CF). Multiple logistic regression showed that both phenotypes are predicted by the level of deprivation before infection (adjusted p-value for CF and AD-CF = 0.025 and 0.0054 respectively). While the severity of acute COVID (adjusted p-value = 0.023) and the number of pre-existing medical conditions predict only the CF phenotypes (adjusted p-value = 0.003), past suicidal ideas predict the AD-CF phenotype (adjusted p-value < 0.001). Pre-infection vaccination status did not predict either phenotype. Our finding suggests that we should recognize the heterogeneity under the umbrella of chronic PCNPS, and a holistic bio-psycho-social approach is essential in understanding them.

## Introduction

The COVID-19 pandemic has left a lasting impact on people’s lives around the world. Despite the world having been keen to move on from the shadow of the pandemic, there are still many unsolved questions about the long-term consequences of COVID in some of the patients. Neuropsychiatric symptoms, such as fatigue, cognitive impairments, anxiety and depression, are among the most common post-COVID symptoms [1]. These symptoms could be unremitting. lasting for over two years after infection [2]. An elevated incidence rate of psychiatric disorder, such as depression and anxiety disorder, is found among those who recovered from COVID-19 [3–5] which can cause an increase in burden on the mental health system. However, it is challenging to have a full comprehension of chronic post-COVID neuropsychiatric symptoms (PCNPS) due to various factors including variants of SARS-CoV-2 strain [6], vaccination status [7] and the use of antiviral drugs [8]. Early studies primarily focused on those infected by pre-Omicron strains and hospitalized patients instead of mild acute COVID patients While some studies suggests that the prevalence of post-COVID symptoms among patients infected by the Omicron variant is lower than some of the earlier variants, other studies disagree [9–11]. Afterall, there are still significant knowledge gaps about chronic PCNPS after the Omicron or post-Omicron strains infection. For example, the heterogeneity of chronic PCNPS has not been adequately explored in patients infected by Omicron and post-Omicron variants. While PCNPS are often grouped into one single entity, our previous study suggested that chronic PCNPS among those infected by pre-Omicron strains can be clustered into an anxiety-depression cluster and a cognitive complaint-fatigue cluster [12]. Discovering scientifically meaningful symptom clusters and phenotypes among patients is the basis of further discovery of the phenomenon’s epidemiology, pathophysiology, and treatments. It would be of interest to investigate the heterogeneity of chronic PCNPS among Omicron and post-Omicron strains patients and to see if it follows a similar pattern to that of pre-Omicron patients. In addition, few studies looking into risk factors of chronic PCNPS consider biological factors together with socioeconomic factors, while they potentially confound each other.

The primary aim of our study was to explore the relationship among the chronic PCNPS who suffered from the infection during the ‘Omicron wave’ in Hong Kong in 2022. The secondary aim of the study was to explore the phenotypes among patients suffering from chronic PCNPS, and to identify their distinct clinical trajectories and risk factors. The key hypotheses of our study are 1. chronic PCNPS in patients infected with SARS-CoV-2 over a year ago during the ‘Omicron wave’ showed a similar clustering pattern with symptoms in patients infected with pre-Omicron strain (which is derived from our previous study); 2. These chronic PCNPS are associated with a) clinical risk factors, such as, severity of the acute infection; b) socioeconomic status e.g., level of deprivation; and c) pre-infection vaccination status.

## Methods

The current study is part of the ‘Long-term mental and brain health effects of COVID-19 from the Omicron strains among adult patients’ study. The key hypotheses and outcome measures were pre-registered (https://doi.org/10.1186/ISRCTN11876145). We recruited participants from the community in Hong Kong via online advertisement. The inclusion criteria are: (i) self-report history of a first PCR or rapid antigen test-confirmed SARS-CoV-2 infection that occurred after January 2022 (i.e., After the Omicron variant became dominant in Hong Kong [13]); (ii) the infection occurred at least one year prior to the study, iii) age between 18-65 years. Consented participants underwent a panel of app-based assessment, which include 1) detailed questionnaire on demographic information and socioeconomic and health status at two time points: 12/2021 and recruitment; 2) a COVID symptoms checklist that comprised 16 neuropsychiatric items and 26 non-neuropsychiatric items; 3) standardized mental health, sleep and health-related quality of life (HRQoL) measures; 4) a seven-day sleep diary; and 5) app-based cognitive tasks focusing on key domains (concentration, psychomotor speed and working memory) related to the phenomenon of ‘brain fog’ (see Supplementary material). The study was approved by the Joint CUHK NTEC Clinical Research Ethic committee (Ref. no.: 2022.362) and the Central Hospital Authority Institutional Review Board (Ref. no.: CIRB-2022-006-1).

### Data analysis

Similar to our previous paper, we built a regularized partial correlation network of self-reported chronic PCNPS to explore their relationships and clustering patterns [14, 15]. To minimize the instability of the network, we excluded symptoms suffered by fewer than 1% of the participants. After network estimation, we used the walktrap algorithm to discover the clusters within the symptom network. Steps, the length of the random walks done by the walktrap algorithm. [16]. We used the community assortativity (R_com_) metric, a bootstrapping procedure, to measure the robustness of community assignment done by the walktrap algorithm. Community assignments are deemed to be robust if R_com_ is larger than 0.5 [17]. We used R packages of bootnet, IsingFit, igraph, ggraph and plyr to perform the network estimation process, and asnipe and assortnet for assortativity metric estimation. According to our previous paper, patients with fewer than four chronic PCNPS have a similar level of mental distress as normal control [12]. In the current study, therefore, we first defined a low symptom load group (LL) using this cut-off. We then used the Bernoulli Mixture Model (BMM) to explore the phenotypes among subjects with four or more chronic PCNPS based on their symptoms profile. We ran the BMM fitting algorithm through cluster numbers from 1 to 10, and for each cluster number, we ran a thousand repetitions, and we selected the best fitting model based on Bayesian information criterion (BIC). We used the flexmix R package to fit the BMM [18].

We tested for univariate between-group differences in demographics, socioeconomic and health status, and symptom score between the subgroups using non-parametric statistical tests. If the variable was categorical, Pearson’s chi-squared test was used; if there was significant statistical difference among the groups, Pearson’s chi-squared test was also used to conduct the post-hoc pairwise comparison tests, but the Bonferroni correction was applied onto the test results to counteract the multiple comparisons problem. Same methodology was applied to ordinal and numerical variables, except that Kruskal-Wallis H Test and Dunn’s test (post-hoc) were used. We used multinomial logistic regression to predict participants’ membership of phenotypes discovered by the BMM model, with the LL group as the reference group.

## Results

We recruited 1273 subjects, and 1205 completed the assessment. The mean age of the subjects was 38.8 and 54.9% were female. 2.2 % of the sample are of non-Chinese ethnicity. The subjects had their first SARS-CoV-2 infection from January 2, 2022, to September 10, 2023, and 24.6% had moderate to severe acute COVID (defined as having symptoms of pneumonia and/or requiring oxygen therapy). Prior to the index infection, 15.0% and 5.4% of them had at least one medical and psychiatric comorbidities, respectively.

### Network analysis

The most frequently reported chronic PCNPS in our cohort are memory problems (39.2%), inability to concentrate (23.9%), fatigue (21.8%), insomnia (14.7%) and post-traumatic stress (PTS) (12.5%, as measured by IES-R) (Table 1), which are similar to our pre-Omicron cohort [12]. Our partial correlation network analysis discovered two major symptom clusters (the cognitive complaint-fatigue cluster and anxiety-depression cluster) and a minor cluster (headache-dizziness cluster) (Figure 1). This clustering pattern and the cluster membership are very similar to our previous results from the pre-Omicron cohort [12].

**Figure 1.**
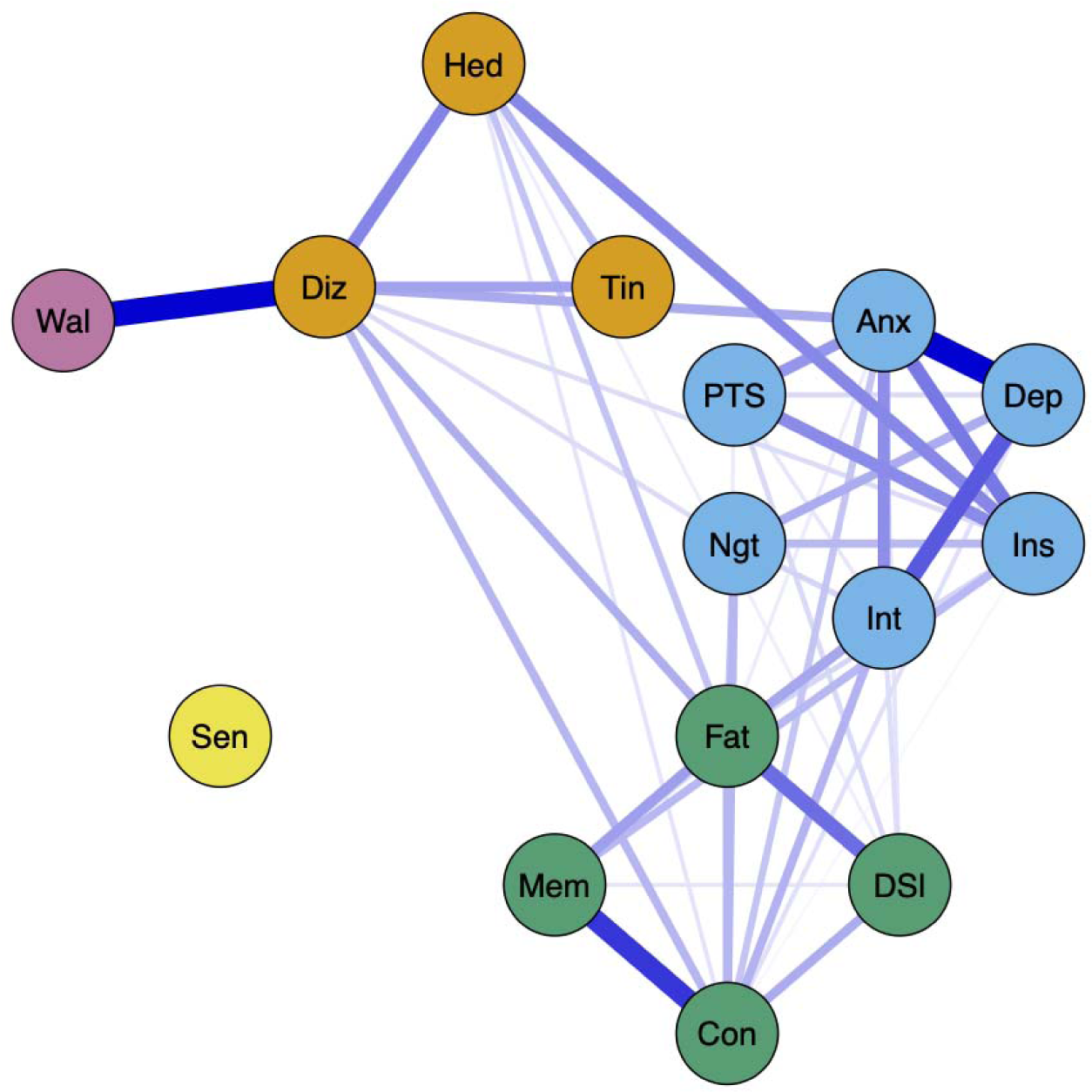
Partial correlation network of post-COVID neuropsychiatric symptoms. *Note. Fat* : fatigue, *Con* : inability to concentrate, *Mem* : memory problems, *DSl* : daytime sleepiness, *Anx* : feeling anxious, *Dep* : feeling depressed, *Int* : loss of interest or pleasure, *PTS* : COVID-related post-traumatic stress symptoms, *Ins* : insomnia, *Ngt* : frequent nightmare, *Hed* : headache, *Diz* : dizziness, *Tin* : tinnitus, *Wal* : imbalanced walking, *Sen* : loss or change to your sense of taste and smell. The colour of the node represents the cluster they belong to.

**Table 1.**
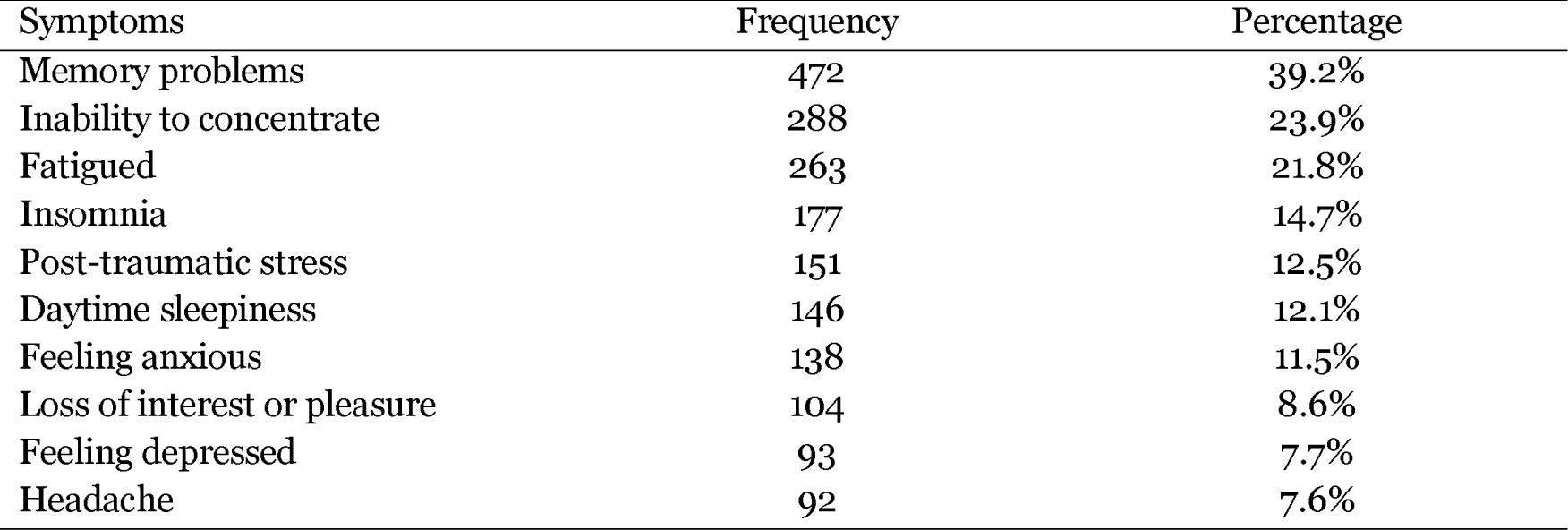
List of ten most frequently reported chronic post-COVID neuropsychiatric symptoms (N=1205)

### Phenotyping using BMM model

The best BMM model resulted in 2 phenotypes among those with high symptom load, namely a type with predominantly cognitive complaints-fatigue cluster symptoms (the CF phenotype, n=161) and another type with a high number of symptoms across the anxiety-depressive cluster and cognitive complaints-fatigue cluster symptoms (the AD-CF phenotype, n=75) (Figure 2).

**Figure 2.**
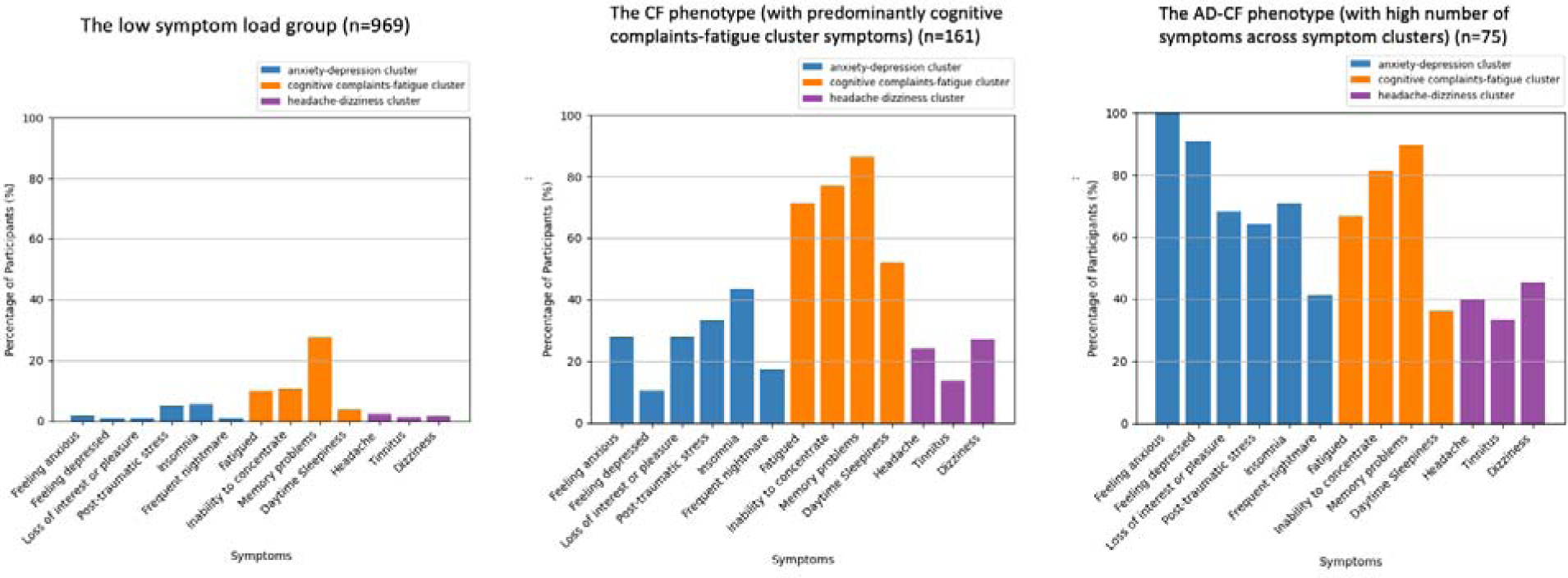
The symptom profile of the low symptom load group, the CF phenotype and the AD-CF phenotype.

### Univariate comparisons across the LL, CF and AD-CF groups

Comparing the LL, CF and AD-CF groups, their age and sex ratio are similar. The LL group had the least number of medical illnesses (mean LL vs CF vs AD-CF = 0.2 vs 0.4 vs 0.3, p<0.001; post-hoc test LL < CF, LL < AD-CF) and psychiatric illnesses (LL vs CF vs AD-CF = 4% vs 10% vs 11%, p = 0.0014; post-hoc test LL < CF, LL < AD-CF), and they were the least deprived (as measured by the Deprivation Index [19]) before the infection (median LL vs CF vs AD-CF = 0 vs 1 vs 2, p<0.001; post-hoc test LL < CF, LL < AD-CF) (Table 2a). The LL group also had the least subjects who suffered from moderate to severe acute COVID (LL vs CF vs AD-CF = 22% vs 35% vs 37%, p<0.001; post-hoc test LL < CF, LL < AD-CF) and they had the shortest total duration of pandemic-related home quarantine (median LL vs CF vs AD-CF = 7 vs 10 vs 11, p<0.001; post-hoc test LL < CF, LL < AD-CF). The LL group has the least post-infection emergent physical and mental health problems. In contrast, the CF and AD-CF groups had significantly more post-infection diagnosed psychiatric illnesses (LL vs CF vs AD-CF = 1% vs 4% vs 8%, p<0.001; Post-hoc test LL < CF, LL < AD-CF) and new suicidal idea (LL vs CF vs AD-CF = 3% vs 9% vs 17%, p<0.001; post-hoc test LL < CF, LL < AD-CF) compared to the LL group, and the CF group had more newly diagnosed medical problems (mean LL vs CF vs AD-CF = 0.06 vs 0.16 vs 0.11, p = 0.00027; post-hoc test LL < CF). The AD-CF group had a significant increase in deterioration in socioeconomic status compared to the LL group with an increase in deprivation (mean LL vs CF vs AD-CF = −0.2 vs 0.1 vs 0.6, p = 0.0005; post-hoc test LL < AD-CF) and domestic violence (LL vs CF vs AD-CF = 1% vs 2% vs 4%, p = 0.018; post-hoc test LL < AD-CF)(Table 2b).

**Table 2a.**
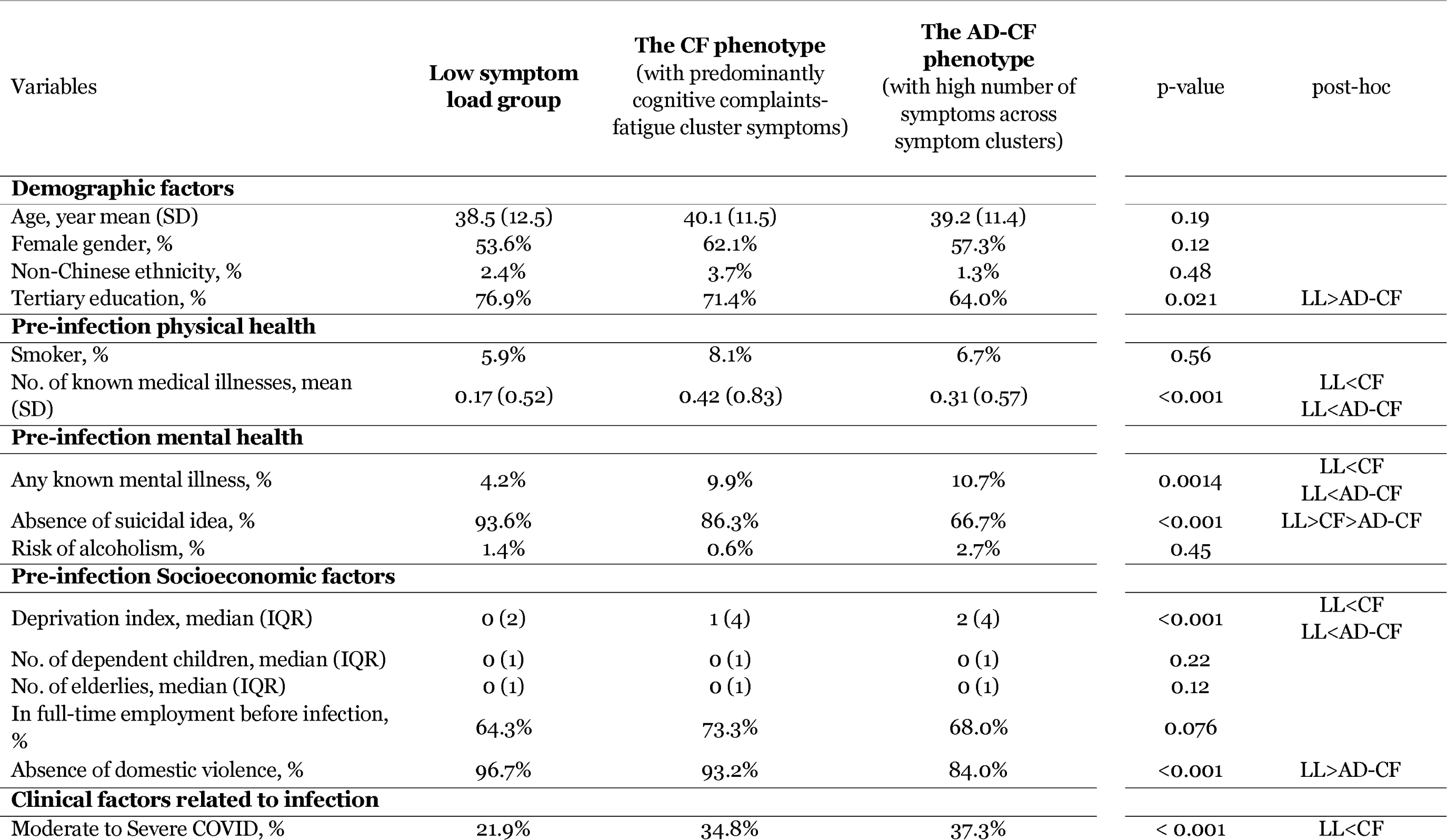
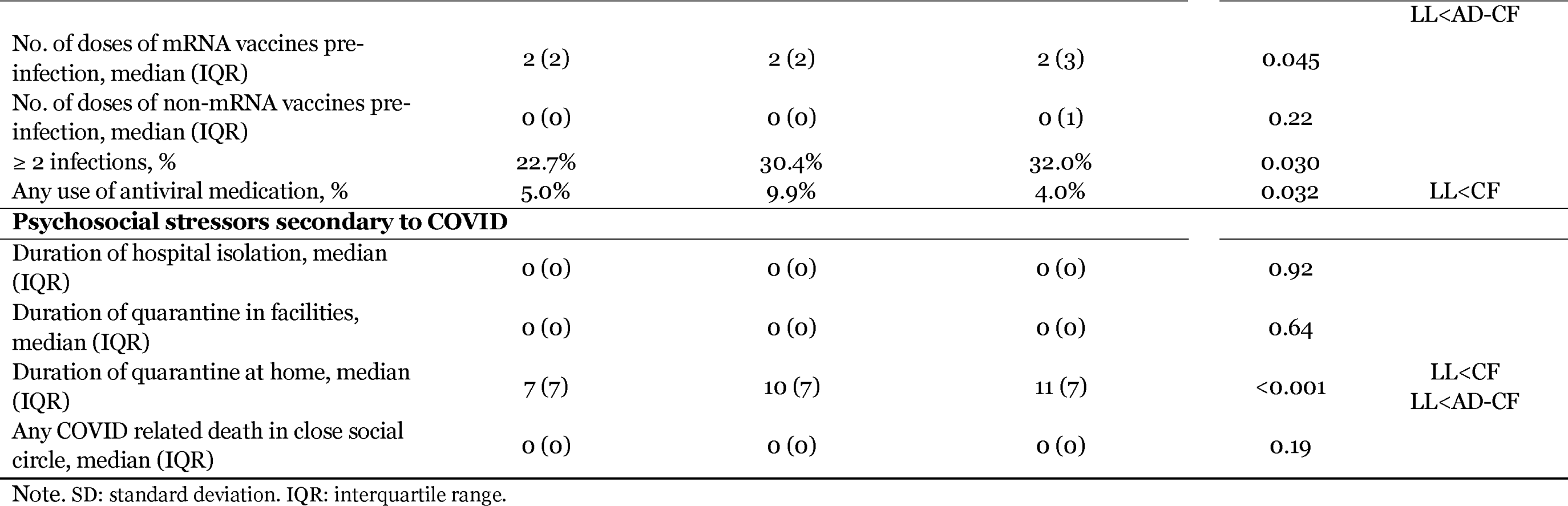
Comparisons of demographic factors, pre-infection physical health mental health and socioeconomic factors, clinical factors related to infection and psychosocial stressors secondary to COVID among the low symptom load group, CF phenotype and AD-CF phenotype.

**Table 2b.** Comparisons of post-infection changes in physical health, mental health, sleep health and socioeconomic factors among the low symptom load group, CF phenotype and AD-CF phenotype.

| Variables | Low symptom load group | The CF phenotype (with predominantly cognitive complaints-fatigue cluster symptoms) | The AD-CF phenotype (with high number of symptoms across symptom clusters) | p-value | post-hoc |
| --- | --- | --- | --- | --- | --- |
| **Post-infection physical health** |  |  |  |  |  |
| No. of new medical diagnoses, mean (SD) | 0.058 (0.25) | 0.16 (0.45) | 0.11 (0.31) | <0.001 | LL<CF |
| No. of days of hospitalization in the past one year, mean (SD) | 0.33 (2.4) | 0.57 (3.2) | 0.48 (1.4) | 0.045 | LL<AD-CF |
| No. of medical consultations in the past one year, mean (SD) | 4.6 (6.9) | 8.4 (9.8) | 8.8 (12.6) | <0.001 | LL<CF<br>LL<AD-CF |
| BMI>30, % | 4.9% | 9.3% | 4.0% | 0.058 |  |
| **Post-infection changes in mental health** |  |  |  |  |  |
| Any newly diagnosed psychiatric illness, % | 0.8% | 3.7% | 8.0% | <0.001 | LL<CF<br>LL<AD-CF |
| New suicidal idea, % | 2.6% | 8.7% | 17.3% | <0.001 | LL<CF<br>LL<AD-CF |
| **Post-infection sleep health** |  |  |  |  |  |
| Pre-infection perceived sleep demand, hour median (IQR) | 8.0 (1.0) | 8.0 (1.0) | 8.0 (1.0) | 0.72 |  |
| Post-infection sleep demand, hour mean (SD) | 8.01 (1.45) | 8.38 (1.33) | 8.55 (2.18) | <0.001 | LL<CF<br>LL<AD-CF |
| Actual average sleep time, hour median (IQR) | 6.94 (1.36) | 6.33 (1.68) | 6.15 (1.58) | <0.001 | LL>CF<br>LL>AD-CF |
| Sleep latency, hour median (IQR) | 0.17 (0.17) | 0.25 (0.42) | 0.33 (0.5) | <0.001 | LL<CF<br>LL<AD-CF |
| Mid-sleep arousal, hour median (IQR) | 0.083 (0.17) | 0.13 (0.33) | 0.21 (0.48) | <0.001 | LL<CF<br>LL<AD-CF |
| **Post-infection changes in socioeconomic factors** |  |  |  |  |  |
| Changes in deprivation index, mean (SD) | -0.168 (1.60) | 0.143 (2.18) | 0.64 (3.85) | <0.001 | LL<AD-CF |
| Any new domestic violence, % | 0.7% | 1.9% | 4.0% | 0.018 | LL<AD-CF |
| Currently in full-time employment, % | 18.1% | 26.7% | 24.0% | 0.023 | LL<CF |

Note. SD: standard deviation, IQR: interquartile range.

**Table 2c.**
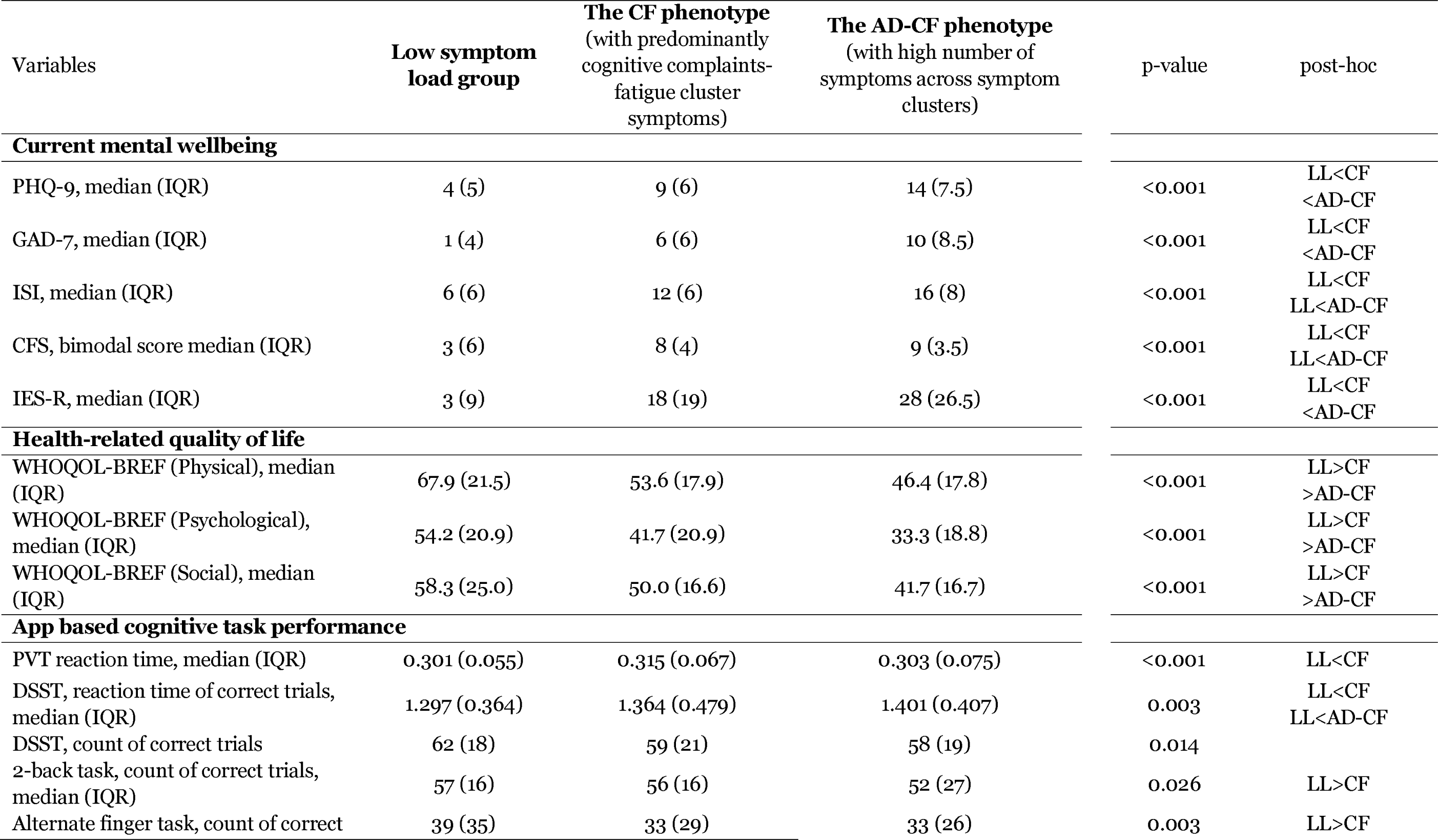
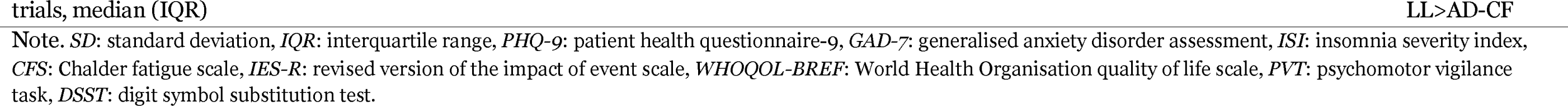
Comparisons of current mental wellbeing, health-related quality of life and app based cognitive task performance among low symptom load group, CF phenotype and AD-CF phenotype.

For their mental and physical health at the time of the assessment, the LL group had the least self-reported mental health distress across all measures, and they enjoyed the best HRQoL (Table 2c). The AD-CF group scored the worst in most of the mental health and HRQoL measures, but was not significantly worse than the CF group in terms of level of insomnia (ISI median LL vs CF vs AD-CF = 6 vs 12 vs 16, p<0.001; post hoc test LL < CF, LL < AD-CF) and fatigue (CFQ median LL vs CF vs AD-CF = 3 vs 8 vs 9, p<0.001; post hoc test LL < CF, LL < AD-CF). The CF and AD-CF group have higher current sleep demand than the LL group (hours, mean LL vs CF vs AD-CF = 8.0 vs 8.4 vs 8.5, p<0.001; post hoc test LL < CF, LL < AD-CF), while their pre-infection sleep demand was identical. However, the CF and AD-CF groups have shorter sleep duration (hours, median LL vs CF vs AD-CF = 6.9 vs 6.3 vs 6.1, p<0.001; post-hoc test LL > CF, LL > AD-CF), longer sleep latency (minutes, median LL vs CF vs AD-CF = 10 vs 15 vs 20, p<0.001; post hoc test LL < CF, LL < AD-CF, CF < AD-CF) and more midnight arousal (mean LL vs CF vs AD-CF = 0.8 vs 1.2 vs 1.6, p<0.001; post hoc test LL < CF, LL < AD-CF) than the LL group as they reported in sleep diary (Table 2c).

The CF group performed worse than LL group in the persistent vigilance test (PVT) (ms, mean LL vs CF vs AD-CF = 0.4 vs 0.5 vs 0.4, p = 0.00042; post hoc test LL< CF) and the digit symbol substitution test (DSST) reaction time (ms, median LL vs CF vs AD-CF =1.3 vs 1.4 vs 1.4, p = 0.0029; post hoc test LL < CF, LL < AD-CF), and alternate finger tapping (correct count, median LL vs CF vs AD-CF = 39 vs 33 vs 33, p = 0.0028; post hoc test LL > CF, LL > AD-CF), while the AD-CF group performed worse in DSST reaction time, 2 back task (correct count, median LL vs CF vs AD-CF = 57 vs 56 vs 52, p = 0.026; post hoc test LL > AD-CF) and alternate finger tapping task (correct count, median LL vs CF vs AD-CF = 39 vs 33 vs 33, p = 0.0028; post hoc test LL > CF, LL > AD-CF) (Table 2c).

### Shared but also distinct predictors for CF and AD-CF subgroups

Using multiple logistic regression controlling for demographic factors, pre-infection physical and mental health status, pre-infection socioeconomic factors, clinical factors related to index acute COVID and psychosocial stressors due to COVID, we found that pre-infection level of deprivation and total quarantine duration related to the pandemic are predictive of both CF (adjusted odd ratio =1.39, 95% CI [1.10, 1.76], adjusted p = 0.025; and adjusted odd ratio =1.04, 95% CI [1.01, 1.06], adjusted p=0.023, respectively) and AD-CF (adjusted odd ratio =1.76, 95% CI [1.27, 2.43], adjusted p = 0.0054; and adjusted odd ratio =1.05, 95% CI [1.02, 1.08], adjusted p=0.0054, respectively) group status. Number of known medical illness pre-infection (adjusted odd ratio =1.63, 95% CI [1.26, 2.11], adjusted p = 0.003), moderate to severe acute COVID severity (adjusted odd ratio =1.76, 95% CI [1.20, 2.59], adjusted p = 0.023) and having full-time job before the infection (adjusted odd ratio =2.0, 95% CI [1.32, 3.05], adjusted p = 0.01) preferentially predict CF status, while pre-infection suicidal idea predicts AD-CF status only (adjusted odd ratio =0.18, 95% CI [0.09, 0.34], adjusted p<0.001)(Table 3).

**Table 3.** Multinomial logistic regression: predictors of high symptom load phenotypes, with low symptom load group as reference group.

| Variables | The CF phenotype<br>(with predominantly cognitive complaints-fatigue cluster symptoms) |  |  |  | The AD-CF phenotype<br>(with high number of symptoms across symptom clusters) |  |  |  |
| --- | --- | --- | --- | --- | --- | --- | --- | --- |
|  | Adjusted OR | 95% CI |  | Adjusted p-value | Adjusted OR | 95% CI |  | Adjusted p-value |
| constant | 0.06 | 0.02 | 0.26 | 0.0030 | 0.20 | 0.03 | 1.22 | 0.32 |
| **Demographic factors** |  |  |  |  |  |  |  |  |
| Age | 1.00 | 0.98 | 1.02 | 0.93 | 1.01 | 0.98 | 1.04 | 0.67 |
| Female gender | 1.45 | 1.00 | 2.10 | 0.15 | 1.01 | 0.60 | 1.71 | 0.96 |
| Non-Chinese ethnicity | 1.26 | 0.45 | 3.48 | 0.82 | 0.47 | 0.05 | 4.49 | 0.60 |
| Tertiary education | 0.95 | 0.60 | 1.52 | 0.93 | 0.57 | 0.29 | 1.09 | 0.32 |
| BMI > 30 | 1.44 | 0.73 | 2.82 | 0.49 | 0.55 | 0.15 | 1.99 | 0.56 |
| **Pre-infection physical health** |  |  |  |  |  |  |  |  |
| Smoking | 1.15 | 0.57 | 2.30 | 0.82 | 0.73 | 0.26 | 2.07 | 0.60 |
| No. of known medical illness | **1.63** | **1.26** | **2.11** | **0.0030** | 1.40 | 0.93 | 2.11 | 0.35 |
| **Pre-infection mental health** |  |  |  |  |  |  |  |  |
| Known mental illness | 1.88 | 0.95 | 3.73 | 0.18 | 1.36 | 0.54 | 3.42 | 0.60 |
| Absence of suicidal idea | 0.58 | 0.32 | 1.05 | 0.18 | **0.18** | **0.09** | **0.34** | **5.6E-06** |
| Risk of alcoholism | 0.43 | 0.05 | 3.50 | 0.64 | 1.67 | 0.31 | 9.10 | 0.60 |
| **Pre-infection Socioeconomic factors** |  |  |  |  |  |  |  |  |
| Deprivation index (log-transformed) | **1.39** | **1.10** | **1.76** | **0.025** | **1.76** | **1.27** | **2.43** | **0.0054** |
| No. of dependent children | 1.24 | 1.01 | 1.53 | 0.15 | 1.22 | 0.89 | 1.69 | 0.45 |
| Absence of domestic violence | 0.61 | 0.28 | 1.32 | 0.37 | 0.44 | 0.19 | 1.03 | 0.29 |
| No. of elderlies | 1.19 | 0.91 | 1.56 | 0.37 | 1.33 | 0.92 | 1.92 | 0.36 |
| Work full-time pre-infection | **2.00** | **1.32** | **3.05** | **0.010** | 1.51 | 0.84 | 2.72 | 0.42 |
| **Clinical factors** |  |  |  |  |  |  |  |  |
| Moderate to Severe COVID | **1.76** | **1.20** | **2.59** | **0.023** | 2.03 | 1.18 | 3.51 | 0.066 |
| No. of mRNA vaccines pre-infection | 1.00 | 0.82 | 1.22 | 0.98 | 0.85 | 0.65 | 1.11 | 0.45 |
| No. of non-mRNA vaccines pre-infection | 1.17 | 0.93 | 1.48 | 0.36 | 0.88 | 0.64 | 1.23 | 0.60 |
| >=2 infection | 1.19 | 0.79 | 1.77 | 0.63 | 1.31 | 0.74 | 2.31 | 0.56 |
| Use of antiviral medication | 1.76 | 0.90 | 3.47 | 0.23 | 0.65 | 0.18 | 2.40 | 0.60 |
| **Psychosocial stressors secondary to COVID** |  |  |  |  |  |  |  |  |
| Duration of hospital isolation | 0.97 | 0.87 | 1.08 | 0.81 | 0.95 | 0.83 | 1.10 | 0.60 |
| Duration of quarantine in facilities | 1.00 | 0.97 | 1.03 | 0.93 | 1.02 | 0.98 | 1.05 | 0.56 |
| Duration of quarantine at home | **1.04** | **1.01** | **1.06** | **0.023** | **1.05** | **1.02** | **1.08** | **0.0054** |
| No. of COVID related death in close social circle | 1.17 | 0.75 | 1.84 | 0.67 | 0.62 | 0.30 | 1.30 | 0.45 |

## Discussion

To the best of our knowledge, this is the first study using pre-registered protocol to replicate the clustering of chronic PCNPS in cohorts infected by different SARS-CoV-2 virus variants. This is also the first study phenotyping patients infected during the Omicron wave with chronic PCNPS by their symptom profile and to examine their demographical, socioeconomic, health and clinical risk factors.

### Neuropsychiatric symptoms in patients infected with Omicron strains of SARS-CoV-2 over a year ago showed a similar clustering pattern with symptoms in patients infected with pre-Omicron strain

The network analysis result on our ‘Omicron wave’ cohort replicated the clustering pattern we found from our smaller, pre-Omicron cohort, which supported our primary hypothesis-both contain a cognitive complaint-fatigue cluster, a depression-anxiety symptoms cluster and a headache-dizziness cluster with highly comparable cluster membership. In recent years, the failure to replicate results in scientific studies, especially in the field of psychiatry or psychology, has raised concerns [20]. Validation by independent cohorts is a gold standard in ensuring generalisability of findings. Understanding the relationship and groupings among symptoms is fundamental to patient stratification, and subsequently biomarker discovery and interventional study, as heterogeneity within the study population introduce noise and reduces power [21]. For example, one large intervention trial on probiotics for long COVID symptoms was shown to be effective on fatigue and cognitive complaints, but not mood complaints [22].

### Differential natural clinical and socioeconomic trajectories for different phenotypes

The LL, CF and AD-CF groups had very different pre-infection socioeconomic and health status prior to the infection. This discrepancy grew after their infection, with both the CF and AD-CF phenotypes showing the increase in physical and mental health problems. The AD-CF group particularly suffered most socially with increased domestic violence and deprivation. However, our analysis limits us from inferring the causal relationship between post-infection health burden, socioeconomic changes and the presence of chronic PCNPS.

### Pre-existing health vulnerabilities and clinical severity of COVID predicts different phenotypes

Our results echo existing evidence that pre-existing mental health, physical health, and clinical severity of acute COVID predict the presence of chronic post-COVID symptoms, including but not limited to neuropsychiatric symptoms [9, 23, 24]. Yet we are the first study suggesting that different clinical factors predict different chronic PCNPS phenotypes. Patients with predominantly CF cluster symptoms were predicted by physical factors such as pre-existing physical illnesses and severity of acute COVID infection. The clinical severity of acute COVID is shown to be a predictive factor of long COVID in various studies [9, 25, 26]. It is hypothesised that the more severe the initial infection, the more severe immune dysregulation and/or endothelial dysfunction. Pre-existing physical health problems are known to increase risk of more severe COVID [27]. However, how pre-existing health problems contributed to post-COVID neuropsychiatric problem symptoms after adjusting for acute infection severity is not well understood. Interestingly, these two factors do not predict the AD-CF phenotype. On the contrary, the AD-CF phenotype is predicted by pre-infection presence of suicidal idea, a proxy marker of poor mental health. This distinction in risk factors between the two phenotypes suggests that different chronic PCNPS phenotypes might have different underlying mechanisms. Result from a recent longitudinal study find that pre-existing psychiatric problems, but not acute infection severity is predictive of long-term neuropsychiatric outcomes in a group of patients with high prevalence of a mix of post-covid persistent depression, anxiety, fatigue and cognitive impairment, which echoes our findings regarding predictors of the AD-CF phenotype [28].

### Health gap

Deprivation is known to predict negative health outcomes in a wide range of contexts and the public health crisis of COVID pandemic has been no exception: the more deprived populations had higher infection rate initially [29, 30], higher mortality [31], lower healthcare resources accessibility [32], worse psychosocial stress during pandemic [33, 34], and so on. Hastie et al. [35] found that those who had deterioration of health post-COVID in a large UK cohort were more deprived after adjusting for potential confounders. Our previous study showed that level of deprivation before infection predicts higher loading of chronic PCNPS in a separate, pre-Omicron cohort [12], and pre-infection deprivation also predicts both the CF and AD-CF phenotypes in our ‘Omicron wave’ cohort, which supports our hypothesis. This replicable result suggests that social mechanisms of chronic PCNPS should not be overlooked in the quest to understand this phenomenon.

### The vaccination question

Contrary to our hypothesis, we do not find vaccination to be a protective factor against chronic post-COVID neuropsychiatric symptoms. Studies generally agree on the positive protective effect of vaccination against post-COVID symptom [36–38], with a large-scale database study by Lundberg-Morris et al. [39] demonstrated that vaccination is least effective in preventing post-COVID symptoms in patients infected by omicron variant. However, most of these studies use a shorter time frame of 4 weeks to 6 months to define post-COVID symptoms. Some studies on more long-term outcomes (>1 year) show negative results [40]. Our study on the pre-Omicron cohort demonstrated the protective effect of pre-infection covid vaccination on chronic PCNPS, but not in the current ‘Omicron wave’ cohort. Whether this discrepancy is related to the virulence of the different strains, the reduced effectiveness of first-generation COVID vaccine on later strains, the chronicity of the symptoms, or a combination of these factors cannot be ascertained at this point.

### The lasting effect of prolonged quarantine

Previous studies on past pandemics/epidemics showed a relationship between longer quarantine periods and poor mental health outcomes [41]. Multiple studies showed quarantine is associated with an increase in mental health distress during the COVID pandemic [42, 43]. We are the first study that reports an association between chronic PCNPS beyond one year and duration of quarantine during the pandemic. Quarantine affects mental health by increasing social isolation and disruption of daily routine. In Hong Kong where living space is limited, home quarantine can be particularly stressful and can increase conflict within the household. That might partially explain why isolation in hospitals or facilities does not have the same effect, but further studies will be needed to test this hypothesis.

### Conclusion

Recognising the complexity of chronic PCNPS Our results further confirm that the phenomenon of chronic PCNPS is heterogeneous, which may partially explain some inconsistent results from the literature. Specifically, there is a need to recognise the different phenotypes within the umbrella of chronic PCNPS as they have different clinical and risk factors profile. Further research that investigates the pathophysiology and intervention of chronic PCNPS should take into account the presence of different phenotypes. In addition to biological factors, we need more research on the role of psychosocial mechanisms of chronic PCNPS. Afterall, appreciating the complexity of chronic PCNPS is necessary for us to better identify at-risk patients and deliver effective intervention.

### Strengths and limitations

The major strength of this study is that it is a registered report with pre-specified hypotheses and assessment methods. Furthermore, this is partly a replication of another study on a different cohort, which can demonstrate the generalisability of the results. Statistically we used multiple testing corrections on our multiple regression analysis results, which can reduce false positivity. The study’s major limitation is its convenient sampling method, which can introduce sampling bias. We also relied on self-reported infection status to determine if the subjects belonged to the ‘Omicron wave’ cohort, which is the key eligibility criteria, which cannot rule out false reporting. We also do not have access to the Our assessment relied mostly on self-report measures. In addition, the app-based cognitive test covers key measures focusing on ‘brain fog’ phenomenon but does not cover all cognitive domains. Finally, this study is a cross-sectional design, and the data on subjects’ pre-infection and infection status are subjected to recall bias. Such design also limits our ability to infer causal relationship between predictors and target outcome.

## Acknowledgment

We thank Ms. Ling Tsui for her logistical support in conducting the research.

## Financial support

This work was supported by the Research Grant Council (SWHC, grant number C4061-21G) and the Health Bureau, Hong Kong SAR (RHYC and YKW, grant number COVID1903002).

## Declaration of Interest

JWYC received personal fees from Eisai Co., Ltd and travel support from Lundbeck HK limited for overseas conference. CKCL received honoraria for lectures from GSK, and support for attending meetings from Pfizer. YKW received personal fees from Eisai Co., Ltd., for delivering a lecture, and sponsorship from Lundbeck HK Ltd and Aculys Pharma, Inc.

## Transparency Declaration

The lead author and manuscript guarantor (SWHC) affirm that the manuscript is an honest, accurate, and transparent account of the study being reported; that no important aspects of the study have been omitted; and that any discrepancies from the study as planned have been explained.

## Author contribution

SWHC, RNYC, PWCW, and WKW contributed to the design of the study. SWHC, RNYC, PWCW, SXL, YL, JWYC, PKSC, CKCL, TWHL, YKW contributed to the funding acquisition. SWHC, SXL, RNYC and YL contributed to the design of assessment material. SWHC, TMC, TCL and YLL performed data analysis and wrote the first draft of the manuscript. All authors critically reviewed and approved the final manuscript.

## Data availability

The data that support the findings of this study will be openly available after the paper is published

## Notes

### Funding Statement

This study was funded by the Research Grant Council (SWHC, grant number C4061-21G) and the Health Bureau, Hong Kong SAR (RHYC and YKW, grant number COVID1903002).

### Author Declarations

The Clinical Research Ethics Committee of the Joint Chinese University of Hong Kong (CUHK) Hospital Authority New Territories East Cluster (NTEC) (Ref. no.: 2022.362) gave ethical approval for this work. The Central Institution Review Board of the Hospital Authority gave ethical approval for this work (Ref. no.: CIRB-2022-006-1).

