## Supplementary Material for "Chronic post-COVID neuropsychiatric symptoms (PCNPS) persisting beyond one year from being infected during the ‘Omicron wave’"

*Data Analysis*

Network estimation

Similar to our previous paper, we built a regularized partial correlation network of self-reported chronic post-COVID neuropsychiatric symptoms to explore their relationships and clustering patterns. Each node (variable) represented a symptom (such as fatigue, headache, daytime sleepiness); each edge between nodes represented the regularized partial correlation between the symptoms, controlling for the effects of other symptoms in the network model. To minimize the instability of the network, we excluded symptoms suffered by fewer than 1% of the participants. As the model fully consisted of binary variables, an Ising model was used to estimate the network from the symptoms data, with LASSO regularization that reduced spurious partial correlations / edges and resulted in a more parsimonious model. When estimating the network model, a number of network models was being estimated under different values of the LASSO’s tuning parameter lambda. Subsequently, we selected the model with the lowest Extended Bayesian Information Criterion. After network estimation, we used the *walktrap* algorithm, a community detection algorithm, to discover the clusters within the symptom network. *Steps*, the length of the random walks done by the *walktrap* algorithm, was set to be 2. As the walktrap algorithm cannot handle negative edge values, we reset those edges to a value of 0.

Validation of community detection

To validate the robustness of the detected communities, we used a metric called community assortativity (R_com_) to measure the robustness of community assignment done by the walktrap algorithm. To calculate community assortativity, we first generated 1000 bootstrap replicate networks (networks estimated from re-sampling the symptoms data **with** replacement); among these 1000 replicate networks, we measured “the degree to which pairs assigned to the same community in the empirical network also occur in the same community in bootstrap replicate networks”. If a network’s R_com_ is equal to 1, it implies the highest robustness of community assignment, where community assignments in all bootstrap replicate networks were identical to those in the empirical network. This also indicates that the network likely contains discrete clusters. If R_com_ is equal to 0, it implies that community assignments were essentially random with regard to the original community assignments in the empirical network. The community assignments are deemed to be robust if R_com_ is larger than 0.5.

The R_com_ of the network generated based on our symptoms data was 0.55, indicating our network model likely contained discrete clusters and the walktrap algorithm reliably detected those clusters / communities. Community assortativity was calculated using adapted code provided by Shizuka & Farine (2016).

We used R packages of *bootnet*, *IsingFit*, *igraph*, *ggraph* and *plyr* to perform the network estimation process, and *asnipe* and *assortnet* for assortativity metric estimation.

Phenotyping participants with four or more chronic PCNPS

According to our previous paper, patients with fewer than four chronic PCNPS have a similar level of mental distress as normal control. In the current study, therefore, we first defined a *low symptom load* group using this cut-off. We then used the Bernoulli Mixture Model (BMM) to explore the phenotypes among those with four or more chronic PCNPS. We ran the BMM fitting algorithm through cluster numbers from 1 to 10, and for each cluster number, we ran a thousand repetitions, and we selected the best fitting model based on Bayesian information criterion (BIC). We used the *flexmix* R package to fit the BMM.

Univariate between-group differences

We tested for univariate between-group differences in demographics, socioeconomic and health status, and symptom score between the subgroups using non-parametric statistical tests. If the variable was categorical, Pearson’s chi-squared test was used; if there was significant statistical difference among the groups, Pearson’s chi-squared test was also used to conduct the post-hoc pairwise comparison tests, but the Bonferroni correction was applied onto the test results to counteract the multiple comparisons problem. Same methodology was applied to ordinal and numerical variables, except that Kruskal-Wallis H Test and Dunn’s test (post-hoc) were used. We used multinomial logistic regression to predict participants’ membership in the 3 groups, with the *low symptom load* group as the reference group.

*Participant Recruitment Flowchart*

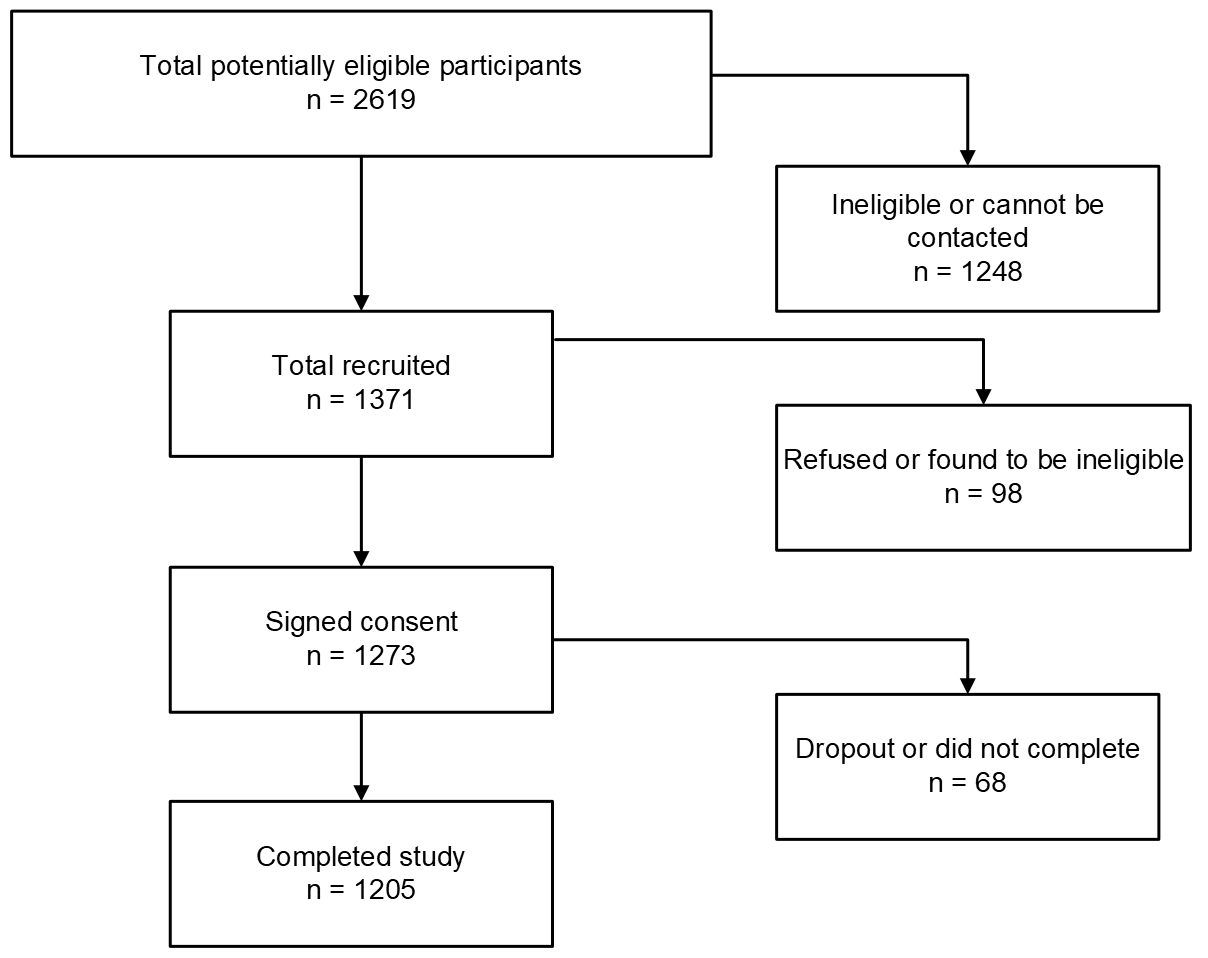

| *Cognitive Test Instructions* | |
| --- | --- |
| **Reaction Test** | |
|  | 1: Please pay attention to the black box at the center of the screen.  2: When a red number appears in the box, click anywhere on the screen as soon as possible.  3: Avoid clicking on the screen before the numbers appear. |
| **Number Symbol Test** | |
|  | 1: A symbol will appear in the center of the screen.  2: Please find the number corresponding to this symbol from the blue table at the top of the screen.  3: And click the orange button with the corresponding number at the bottom of the screen. |
| **Memory Test** | |
| **1-Back memory test** | |
|  | 1: In this test, different English letters will be presented in sequence. If the current letter presented is identical to the previous (1) letter, click “Yes”, otherwise click “No”. Please do this as quickly and accurately as possible. |
|  | 2: In this test, a sequence of letters will be presented one after another. When the current letter is presented, you need to judge whether it is the same as the previous (1) letter, if it is the same, click "Yes"; if it is different, click “No”. In the following example, [M, H, F, F] will be presented one by one in order. You need to click “No” when the first three letters [M, H, F] are presented. And when you see the fourth letter [F], you need to click “Yes” because this letter is the same as the previous (1) letter. |
| **2-Back memory test** | |
|  | 1: In this test, different English letters will be presented in sequence. If the current letter presented is identical to the previous (2) letters, click “Yes”, otherwise click “No”. Please do this as quickly and accurately as possible. |
|  | 2: In this test, a sequence of letters will be presented one after another. When the current letter is presented, you need to judge whether it is the same as the previous (2) letters, if it is the same, click “Yes”; if it is different, click “No”. In the following example, [M, H, F, F] will be presented one by one in order. You need to click “No” when the first three letters [M, H, F] are presented. And when you see the fourth letter [F], you need to click “Yes” because this letter is the same as the previous (2) letters. |
| **Finger Reaction and Coordination Assessment** | |
|  | 1: Please use your index and middle fingers to click the pink button on the screen alternately within 10-seconds test time. Each pink button click scores 1 point.  Please click with your index finger first (1)  Click again with your middle finger (2) |

| *List of Post-acute COVID syndrome (PACS) Checklist* | |
| --- | --- |
| **16 neuropsychiatric items** | |
|  | Memory problems |
|  | Inability to concentrate |
|  | Fatigued |
|  | Insomnia |
|  | Daytime Sleepiness |
|  | Headache |
|  | Feeling anxious |
|  | Dizziness |
|  | Tinnitus |
|  | Frequent nightmare |
|  | Loss of interest or pleasure |
|  | Feeling depressed |
|  | Loss or change to your sense of taste and smell |
|  | Imbalanced walking |
|  | Auditory or visual hallucinations |
|  | Loss of hearing |
| **25 non-neuropsychiatric items** | |
|  | Loss of hair |
|  | Menstrual irregularity or pain |
|  | Rashes |
|  | Congested or stuffy nose |
|  | Blurred Vision |
|  | Shortness of breath or trouble breathing |
|  | Cough |
|  | Heart palpitations and /or irregular heartbeat |
|  | Night sweats |
|  | Joint pain |
|  | Abnormal skin sensation e.g. pin and needle sensation or numbness |
|  | Muscle pain |
|  | Constipation |
|  | Sneeze |
|  | Runny or dripping nose |
|  | Chest congestion and /or Chest pain |
|  | Bodily weaknesses |
|  | Abdominal pain and/or bloating and/or heartburn |
|  | Diarrhea and/or nausea or vomiting |
|  | Conjunctivitis |
|  | Sore or painful throat |
|  | Red, swollen and/or strange patches on the tongue |
|  | Tremor |
|  | Loss of appetite |
|  | Fever/High temperature |
| *Note.* Although we include post-traumatic stress (PTS) in our network analysis and symptom clustering as one of the neuropsychiatric symptoms this symptom is measured using the Impact of Event Scale-Revised (IES-R) questionnaire with a score of 24 or above.  However, it is not included in the original PACS symptom checklist. | |

*Comparison of* ***pre-COVID*** *medical diagnoses among low symptom load group, cognitive complaints-fatigue predominant phenotype and anxiety-depression cluster and cognitive complaints-fatigue cluster phenotype.*

| Medical Diagnoses | Low symptom load group (LL) | Cognitive complaints-fatigue predominant phenotype (CF) | Anxiety-depression cluster and cognitive complaints-fatigue cluster phenotype (AD-CF) |
| --- | --- | --- | --- |
|  | n (%) | n (%) | n (%) |
| None | 850 (87.7) | 118 (73.3) | 56 (74.7) |
| Hypertension | 58 (6.0) | 18 (11.2) | 4 (5.3) |
| Chronic (long-term) respiratory diseases, such as asthma, chronic obstructive pulmonary disease (COPD), emphysema or bronchitis | 11 (1.1) | 8 (5.0) | 2 (2.7) |
| Chronic heart disease such as heart failure | 4 (0.4) | 2 (1.2) | 1 (1.3) |
| Chronic kidney disease | 4 (0.4) | 0 (0.0) | 0 (0.0) |
| Chronic liver disease, such as hepatitis | 11 (1.1) | 3 (1.9) | 0 (0.0) |
| Chronic gastrointestinal disease, such as peptic ulcer | 7 (0.7) | 3 (1.9) | 3 (4.0) |
| Major or minor stroke | 2 (0.2) | 2 (1.2) | 1 (1.3) |
| Chronic neurological conditions (except stroke), such as Parkinson’s disease, motor neuron disease, multiple sclerosis (MS), a learning disability or cerebral palsy | 3 (0.3) | 1 (0.6) | 0 (0.0) |
| Diabetes | 19 (2.0) | 6 (3.7) | 1 (1.3) |
| A weakened immune system as the result of conditions such as HIV and AIDS, or immune-suppressing medicines such as steroid tablets or chemotherapy | 3 (0.3) | 1 (0.6) | 0 (0.0) |
| Being seriously overweight (a body mass index (BMI) of 30 or above) | 12 (1.2) | 6 (3.7) | 0 (0.0) |
| Organ transplant recipient | 1 (0.1) | 1 (0.6) | 0 (0.0) |
| Cancer, receiving treatment | 1 (0.1) | 1 (0.6) | 0 (0.0) |
| Cancer, completed treatment | 9 (0.9) | 3 (1.9) | 4 (5.3) |
| Chronic pain | 20 (2.1) | 12 (7.5) | 7 (9.3) |

*Comparison of* ***newly diagnosed*** *medical diagnoses among low symptom load group, cognitive complaints-fatigue predominant phenotype and anxiety-depression cluster and cognitive complaints-fatigue cluster phenotype.*

| Medical Diagnoses | Low symptom load group (LL) | Cognitive complaints-fatigue predominant phenotype (CF) | Anxiety-depression cluster and cognitive complaints-fatigue cluster phenotype (AD-CF) |
| --- | --- | --- | --- |
|  | n (%) | n (%) | n (%) |
| None | 916 (94.5) | 139 (86.3) | 67 (89.3) |
| Hypertension | 11 (1.1) | 3 (1.9) | 0 (0.0) |
| Chronic (long-term) respiratory diseases, such as asthma, chronic obstructive pulmonary disease (COPD), emphysema or bronchitis | 5 (0.5) | 3 (1.9) | 2 (2.7) |
| Chronic heart disease such as heart failure | 0 (0.0) | 1 (0.6) | 0 (0.0) |
| Chronic kidney disease | 1 (0.1) | 0 (0.0) | 0 (0.0) |
| Chronic liver disease, such as hepatitis | 1 (0.1) | 1 (0.6) | 0 (0.0) |
| Chronic gastrointestinal disease, such as peptic ulcer | 2 (0.2) | 2 (1.2) | 0 (0.0) |
| Major or minor stroke | 0 (0.0) | 0 (0.0) | 1 (1.3) |
| Chronic neurological conditions (except stroke), such as Parkinson’s disease, motor neuron disease, multiple sclerosis (MS), a learning disability or cerebral palsy | 2 (0.2) | 1 (0.6) | 0 (0.0) |
| Diabetes | 8 (0.8) | 2 (1.2) | 1 (1.3) |
| A weakened immune system as the result of conditions such as HIV and AIDS, or immune-suppressing medicines such as steroid tablets or chemotherapy | 2 (0.2) | 0 (0.0) | 0 (0.0) |
| Being seriously overweight (a body mass index (BMI) of 30 or above) | 3 (0.3) | 2 (1.2) | 1 (1.3) |
| Organ transplant recipient | 0 (0.0) | 0 (0.0) | 0 (0.0) |
| Cancer, receiving treatment | 1 (0.1) | 0 (0.0) | 0 (0.0) |
| Cancer, completed treatment | 3 (0.3) | 0 (0.0) | 1 (1.3) |
| Chronic pain | 17 (1.8) | 11 (6.8) | 2 (2.7) |

*Comparison of* ***pre-COVID*** *psychiatric diagnoses among low symptom load group, cognitive complaints-fatigue predominant phenotype and anxiety-depression cluster and cognitive complaints-fatigue cluster phenotype.*

| Psychiatric Diagnoses | Low symptom load group (LL) | Cognitive complaints-fatigue predominant phenotype (CF) | Anxiety-depression cluster and cognitive complaints-fatigue phenotype (AD-CF) |
| --- | --- | --- | --- |
|  | n (%) | n (%) | n (%) |
| None | 928 (95.8) | 145 (90.1) | 67 (89.3) |
| Substance use disorder | 0 (0.0) | 0 (0.0) | 0 (0.0) |
| Bipolar disorder | 4 (0.4) | 2 (1.2) | 2 (2.7) |
| Obsessive-compulsive disorder | 0 (0.0) | 1 (0.6) | 1 (1.3) |
| Social phobia | 0 (0.0) | 0 (0.0) | 1 (1.3) |
| Other anxiety disorder | 10 (1.0) | 2 (1.2) | 3 (4.0) |
| Psychotic disorder | 4 (0.4) | 1 (0.6) | 0 (0.0) |
| Depression | 22 (2.3) | 11 (6.8) | 5 (6.7) |
| Generalised anxiety disorder | 3 (0.3) | 1 (0.6) | 1 (1.3) |
| Post-traumatic stress disorder | 2 (0.2) | 0 (0.0) | 2 (2.7) |
| Panic disorder | 2 (0.2) | 2 (1.2) | 1 (1.3) |
| Eating disorder | 1 (0.1) | 0 (0.0) | 0 (0.0) |

*Comparison of* ***newly diagnosed*** *psychiatric diagnoses among low symptom load group, cognitive complaints-fatigue predominant phenotype and anxiety-depression cluster and cognitive complaints-fatigue cluster phenotype.*

| Psychiatric Diagnoses | Low symptom load group (LL) | Cognitive complaints-fatigue predominant phenotype (CF) | Anxiety-depression cluster and cognitive complaints-fatigue phenotype (AD-CF) |
| --- | --- | --- | --- |
|  | n (%) | n (%) | n (%) |
| None | 961 (99.2) | 155 (96.3) | 69 (92.0) |
| Substance use disorder | 1 (0.1) | 0 (0.0) | 0 (0.0) |
| Bipolar disorder | 0 (0.0) | 0 (0.0) | 0 (0.0) |
| Obsessive-compulsive disorder | 0 (0.0) | 0 (0.0) | 0 (0.0) |
| Social phobia | 0 (0.0) | 0 (0.0) | 0 (0.0) |
| Other anxiety disorder | 4 (0.4) | 3 (1.9) | 4 (5.3) |
| Psychotic disorder | 0 (0.0) | 0 (0.0) | 2 (2.7) |
| Depression | 2 (0.2) | 3 (1.9) | 1 (1.3) |
| Generalised anxiety disorder | 0 (0.0) | 1 (0.6) | 0 (0.0) |
| Post-traumatic stress disorder | 0 (0.0) | 1 (0.6) | 0 (0.0) |
| Panic disorder | 1 (0.1) | 0 (0.0) | 0 (0.0) |
| Eating disorder | 0 (0.0) | 0 (0.0) | 0 (0.0) |

| *Frequencies of COVID* ***neuropsychiatric symptoms*** *among low symptom load group, cognitive complaints-fatigue predominant phenotype and anxiety-depression cluster and cognitive complaints-fatigue phenotype.* | | | | | | | | |
| --- | --- | --- | --- | --- | --- | --- | --- | --- |
| Neuropsychiatric Symptoms | Low symptom load group (LL) | |  | Cognitive complaints-fatigue predominant phenotype (CF) | |  | Anxiety-depression cluster and cognitive complaints-fatigue cluster phenotype (AD-CF) | |
|  | Frequency | Percentage |  | Frequency | Percentage |  | Frequency | Percentage |
| Memory problems | 266 | 27.5% |  | 139 | 86.3% |  | 67 | 89.3% |
| Inability to concentrate | 103 | 10.6% |  | 124 | 77.0% |  | 61 | 81.3% |
| Fatigued | 97 | 10.0% |  | 116 | 72.0% |  | 50 | 66.7% |
| Insomnia | 54 | 5.6% |  | 70 | 43.5% |  | 53 | 70.7% |
| Post-traumatic stress | 49 | 5.1% |  | 54 | 33.5% |  | 48 | 64.0% |
| Daytime Sleepiness | 36 | 3.7% |  | 83 | 51.6% |  | 27 | 36.0% |
| Headache | 23 | 2.4% |  | 39 | 24.2% |  | 30 | 40.0% |
| Feeling anxious | 18 | 1.9% |  | 45 | 28.0% |  | 75 | 100.0% |
| Dizziness | 14 | 1.4% |  | 43 | 26.7% |  | 34 | 45.3% |
| Tinnitus | 13 | 1.3% |  | 22 | 13.7% |  | 25 | 33.3% |
| Frequent nightmare | 9 | 0.9% |  | 27 | 16.8% |  | 31 | 41.3% |
| Loss of interest or pleasure | 9 | 0.9% |  | 44 | 27.3% |  | 51 | 68.0% |
| Feeling depressed | 8 | 0.8% |  | 17 | 10.6% |  | 68 | 90.7% |
| Loss or change to your sense of taste and smell | 5 | 0.5% |  | 4 | 2.5% |  | 5 | 6.7% |
| Imbalanced walking | 3 | 0.3% |  | 20 | 12.4% |  | 15 | 20.0% |
| Auditory or visual hallucinations | 0 | 0.0% |  | 4 | 2.5% |  | 5 | 6.7% |
| Loss of hearing | 0 | 0.0% |  | 4 | 2.5% |  | 1 | 1.3% |

| *Frequencies of COVID* ***non-neuropsychiatric symptoms*** *among low symptom load group, cognitive complaints-fatigue predominant phenotype and anxiety-depression cluster and cognitive complaints-fatigue phenotype.* | | | | | | | | |
| --- | --- | --- | --- | --- | --- | --- | --- | --- |
| Non-neuropsychiatric symptoms | Low symptom load group (LL) | |  | Cognitive complaints-fatigue predominant phenotype (CF) | |  | Anxiety-depression cluster and cognitive complaints-fatigue cluster phenotype (AD-CF) | |
|  | Frequency | Percentage |  | Frequency | Percentage |  | Frequency | Percentage |
| Loss of hair | 88 | 9.1% |  | 53 | 32.9% |  | 33 | 44.0% |
| Menstrual irregularity or pain | 36 | 3.7% |  | 29 | 18.0% |  | 12 | 16.0% |
| Rashes | 35 | 3.6% |  | 20 | 12.4% |  | 11 | 14.7% |
| Congested or stuffy nose | 32 | 3.3% |  | 19 | 11.8% |  | 10 | 13.3% |
| Blurred Vision | 31 | 3.2% |  | 33 | 20.5% |  | 30 | 40.0% |
| Shortness of breath or trouble breathing | 31 | 3.2% |  | 37 | 23.0% |  | 27 | 36.0% |
| Cough | 30 | 3.1% |  | 18 | 11.2% |  | 13 | 17.3% |
| Heart palpitations and /or irregular heartbeat | 30 | 3.1% |  | 33 | 20.5% |  | 35 | 46.7% |
| Night sweats | 29 | 3.0% |  | 29 | 18.0% |  | 21 | 28.0% |
| Joint pain | 28 | 2.9% |  | 38 | 23.6% |  | 27 | 36.0% |
| Abnormal skin sensation e.g. pin and needle sensation or numbness | 27 | 2.8% |  | 23 | 14.3% |  | 17 | 22.7% |
| Muscle pain | 24 | 2.5% |  | 37 | 23.0% |  | 26 | 34.7% |
| Constipation | 23 | 2.4% |  | 28 | 17.4% |  | 15 | 20.0% |
| Sneeze | 22 | 2.3% |  | 10 | 6.2% |  | 6 | 8.0% |
| Runny or dripping nose | 19 | 2.0% |  | 9 | 5.6% |  | 8 | 10.7% |
| Chest congestion and /or Chest pain | 17 | 1.8% |  | 22 | 13.7% |  | 19 | 25.3% |
| Bodily weaknesses | 15 | 1.5% |  | 27 | 16.8% |  | 22 | 29.3% |
| Abdominal pain and/or bloating and/or heartburn | 14 | 1.4% |  | 27 | 16.8% |  | 27 | 36.0% |
| Diarrhea and/or nausea or vomiting | 11 | 1.1% |  | 14 | 8.7% |  | 18 | 24.0% |
| Conjunctivitis | 8 | 0.8% |  | 2 | 1.2% |  | 2 | 2.7% |
| Sore or painful throat | 8 | 0.8% |  | 8 | 5.0% |  | 7 | 9.3% |
| Red, swollen and/or strange patches on the tongue | 5 | 0.5% |  | 4 | 2.5% |  | 2 | 2.7% |
| Tremor | 3 | 0.3% |  | 6 | 3.7% |  | 9 | 12.0% |
| Loss of appetite | 3 | 0.3% |  | 7 | 4.3% |  | 8 | 10.7% |
| Fever/High temperature | 1 | 0.1% |  | 4 | 2.5% |  | 2 | 2.7% |
